# Association of extended dosing intervals or delays in pembrolizumab-based regimens with survival outcomes in advanced non-small cell lung cancer

**DOI:** 10.1101/2020.03.31.20048637

**Authors:** Kartik Sehgal, Anushi Bulumulle, Heather Brody, Ritu R. Gill, Shravanti Macherla, Aleksandra Qilleri, Danielle C. McDonald, Cynthia R. Cherry, Meghan Shea, Mark S. Huberman, Paul A. VanderLaan, Glen J. Weiss, Paul R. Walker, Daniel B. Costa, Deepa Rangachari

**Affiliations:** Division of Medical Oncology, Department of Medicine, Beth Israel Deaconess Medical Center, Harvard Medical School, Boston, MA 02215, USA; Division of Hematology and Oncology, Department of Internal Medicine, Brody School of Medicine at East Carolina University, Greenville, NC 27858, USA; Vidant Medical Center, Greenville, NC 27834, USA; Department of Radiology, Beth Israel Deaconess Medical Center, Harvard Medical School, Boston, MA 02215, USA; Department of Pathology, Beth Israel Deaconess Medical Center, Harvard Medical School, Boston, MA 02215, USA

**Keywords:** pembrolizumab, extended dosing intervals, treatment delays, non-small cell lung cancer, patient-physician preference

## Abstract

**Background:** Besides modeling/simulation-based analysis, no post-approval studies have evaluated optimal administration frequency of pembrolizumab in non-small cell lung cancer (NSCLC).

**Patients and Methods:** We performed a multicenter retrospective cohort study to evaluate association between survival outcomes and treatment extensions/delays of pembrolizumab-based regimens in advanced NSCLC patients. Those who had received at least four cycles in routine practice were divided into two groups: non-standard (Non-Std: ≥2 cycles at intervals >3weeks +3days) and standard (Std: all cycles every 3weeks or 1 cycle >3weeks +3days).

**Results:** Among 150 patients, 92 (61%) were eligible for the study (Non-Std:27, Std:65). Reasons for patients with extensions/delays in the Non-Std group included: immune-related adverse events (irAEs,33%), non-irAE-related medical issues (26%), and patient-physician preference (41%). Non-Std group was more likely to have higher PD-L1 tumor proportion score, higher number of treatment cycles and pembrolizumab monotherapy. Univariate and 6-month landmark analyses showed longer median overall survival (OS) and progression-free survival (PFS) in Non-Std group compared to the Std group. After multivariable adjustment for confounding factors, there was no significant difference in OS [HR 1.2 (95%C.I.: 0.3–4.8), p=0.824] or PFS [HR 2.6 (95%C.I.: 0.7–9.6), p=0.157] between the two groups.

**Conclusion:** Our study shows that a significant proportion of advanced NSCLC patients receive pembrolizumab-based regimens with extended intervals or delays in routine clinical practice and with similar outcomes to those receiving treatment at label-specified 3-week intervals. Given the durability of benefit seen and the potential for cost reduction and decreased infusion frequency in these patients, this requires validation in prospective trials.

## INTRODUCTION

The updated results of the KEYNOTE-001 study have confirmed the revolutionary impact of the anti-programmed death-1 (PD-1) agent pembrolizumab on outcomes of patients with advanced non-small cell lung cancer (NSCLC) whose tumors lack actionable oncogenic drivers. ^1-3^ The widespread adoption of anti-PD-1 agents and durable responses seen in some patients have raised important questions regarding the optimal frequency of administration of these drugs, including the impact of treatment interruptions or discontinuations in routine clinical practice. ^4^ Although immune-related adverse events (irAEs) have been associated with improved outcomes in NSCLC ^5,6^, a retrospective study in Canada suggested lower overall survival (OS) in patients receiving interrupted treatments due to irAEs.^7^ Additionally, the lowest and least frequent dose of pembrolizumab that may permit maximal efficacy in advanced NSCLC is still unknown.^4,8^ Moreover, the financial and societal impacts of access to this durably efficacious therapy for this growing population necessitates thoughtful consideration of resource utilization and the patient care experience so as to afford an optimized and sustainable care paradigm for all those who may benefit.^4,9,10^

Recent efforts to develop less frequent and more flexible dosing regimens have included the phase 3b/4 CheckMate 384 study of nivolumab in advanced NSCLC, which confirmed similar efficacy and safety outcomes with 480 mg every 4 weeks compared to 240 mg every 2 weeks, as predicted by exposure-response evaluations.^11,12^ A modeling/simulation study based on the established pharmacokinetic model of pembrolizumab from early developmental trials, predicted that a dose of 400 mg every 6 weeks would be equally as effective as the standard U.S. Food and Drug Administration (FDA)-approved dose of 200 mg every 3 weeks.^13^

However, clinical evaluations of these alternate dosing schemas have not yet been performed. We conducted a multicenter retrospective study to evaluate survival outcomes of patients with advanced NSCLC who were treated with pembrolizumab-based regimens at standard versus extended intervals in routine clinical practice.

## PATIENTS AND METHODS

In this retrospective cohort study, medical charts from 2 tertiary academic cancer centers-Beth Israel Deaconess Medical Center (BIDMC)/ Harvard Medical School and Vidant Medical Center (VMC)/ Brody School of Medicine at East Carolina University - were reviewed in accordance with research protocols approved by the respective institutional review boards. Patients with advanced NSCLC (defined as patients with stage IV or recurrent advanced disease, who were not candidates for curative intent treatment) who received pembrolizumab-based regimens (defined as first-time patients were treated with pembrolizumab in palliative care setting-either as monotherapy or along with chemotherapy) for at least four cycles in routine practice outside clinical trials at either BIDMC or VMC between February 1, 2016 and April 5, 2019 were eligible. Those who started their first pembrolizumab-based regimen outside these two centers were excluded from the study. Patients eligible for the study were divided into two groups: (a) non-standard (Non-Std: those receiving pembrolizumab 200 mg for ≥2 cycles at intervals >3 weeks + 3 days due to any reason), and (b) standard (Std: either all treatment cycles at FDA-approved dose interval or up to 1 cycle at interval >3 weeks + 3 days due to any reason). The objective of this study was to evaluate if advanced NSCLC patients belonging to the Non-Std group had worse OS or progression-free survival (PFS) compared to the Std group.

Patient data was collected on demographics, clinicopathologic characteristics, treatment regimen details, and irAEs. Patient characteristics such as age and ECOG performance status, survival time and duration of response were calculated from the start of first pembrolizumab-based treatment, till progression/ switch to alternative/additional therapy. Tumor molecular profile and mutational burden were evaluated in these patients by different multiplex next-generation sequencing platforms as well as polymerase chain reaction and fluorescence in-situ hybridization for individual mutations/ rearrangements. Disease response was evaluated by thoracic radiologists using the immune Response Evaluation Criteria in Solid Tumors (iRECIST).^14^ Descriptive tables were generated, depicting proportions for categorical variables and median (with range) for non-categorical variables. Fisher exact and Wilcoxon rank sum tests were used to calculate two-sided p values for categorical and continuous outcomes respectively. Kaplan-Meier survival curves and log-rank test was employed for analysis of censored survival outcomes. 6-month landmark analysis was performed to account for immortal time bias. Univariate and multivariable regression to adjust for confounding variables were performed using Cox proportional hazards model. Swimmer’s plot was generated to depict the duration of response from the first non-standard cycle in the Non-Std group. Two-sided p value <0.05 was considered significant. Adjustments for multiple comparisons were not made due to the exploratory nature of this analysis. Graph creation and statistical analysis were performed using Microsoft Excel and Stata/IC v15.1 software.

## RESULTS

Out of 150 patient charts reviewed from both centers, 92 (61%) patients had received at least 4 cycles of pembrolizumab-based regimens and were eligible for the study (**Figure 1**, which demonstrates distribution of screened patients, **and Supplementary Table S1**, which demonstrates characteristics of included and excluded patients). 27 (29%) patients were classified in the Non-Std group, while 65 (71%) belonged to the Std group. Among the Non-Std group patients, 16 had treatment delays due to irAEs (9; 33%) or non-irAE-related medical issues (7, 26%) (**Supplementary Table S2**). 11 (41%) patients opted to receive treatments at extended dosing intervals after a detailed discussion with their physicians. **Table 1** summarizes the patient characteristics of the Non-Std and Std groups. Patients in the Std group were more likely to receive pembrolizumab along with chemotherapy (Non-Std: 29% vs. Std: 66%, p=0.002) and have tumors with lower PD-L1 tumor proportion score (p=0.01). Patients in the Non-Std group were more likely to have higher number of treatment cycles (Non-Std: 14 vs. Std: 6, p<0.0001).

**Figure 1.**
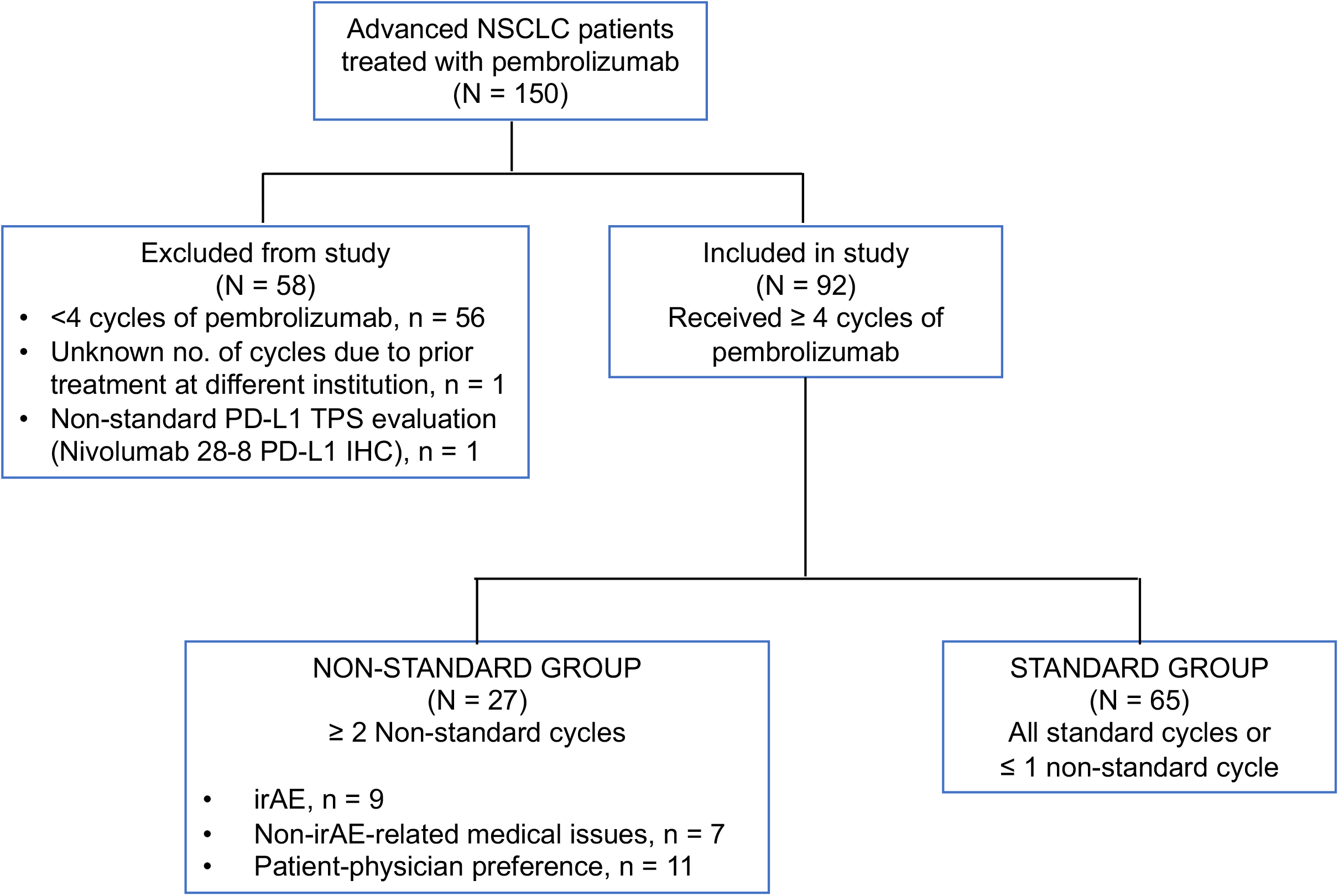
Distribution of patients with advanced NSCLC screened in the study.

**Table 1.** Patient characteristics in the non-standard vs. standard groups

|  | All patients<br>(N = 92) | Standard<br>group<br>(N = 65) | Non-standard<br>group<br>(N = 27) | p |
| --- | --- | --- | --- | --- |
| **Clinico-Pathological Characteristics, n (%) unless specified** |  |  |  |  |
| Median Age, years (range) | 64.5 (37-87) | 64 (49-87) | 66 (37-87) | 0.75 |
| Sex, Female | 44 (48) | 31 (48) | 13 (48) | 1.0 |
| Smoking status, Ever | 84 (91) | 58 (89) | 26 (96) | 0.43 |
| ECOG PS |  |  |  | 0.57 |
| • 0-1 | 75 (82) | 54 (83) | 21 (78) |  |
| • ≥ 2 | 17 (18) | 11 (17) | 6 (22) |  |
| Histology |  |  |  | 1.0 |
| • Non-squamous | 70 (76) | 49 (75) | 21 (78) |  |
| • Squamous | 15 (16) | 11 (17) | 4 (15) |  |
| • Poorly differentiated | 7 (8) | 5 (8) | 2 (7) |  |
| Driver mutation |  |  |  | 0.39 |
| • *KRAS* | 33 (36) | 26 (40) | 7 (26) |  |
| • *EGFR* | 6 (7) | 3 (5) | 3 (11) |  |
| • Others | 3 (3) | 2 (3) | 1 (4) |  |
| • None identified | 40 (43) | 27 (41) | 13 (48) |  |
| • Not assessed | 10 (11) | 7 (11) | 3 (11) |  |
| PD-L1 TPS, % |  |  |  | 0.01 |
| • <1 | 24 (26) | 22 (34) | 2 (7) |  |
| • 1-49 | 17 (18) | 9 (14) | 8 (30) |  |
| • ≥ 50 | 42 (46) | 28 (43) | 14 (52) |  |
| • Not assessed | 9 (10) | 6 (9) | 3 (11) |  |
| TMB, mut/mB |  |  |  | 0.35 |
| • <10 | 20 (22) | 16 (25) | 4 (15) |  |
| • ≥ 10 | 30 (32) | 19 (29) | 11 (41) |  |
| • Not assessed | 42 (46) | 30 (46) | 12 (44) |  |
| **Treatment Characteristics, n (%) unless specified** |  |  |  |  |
| Line of pembrolizumab |  |  |  | 0.05 |
| • First line | 65 (71) | 50 (77) | 15 (56) |  |
| • ≥ Second line | 27 (29) | 15 (23) | 12 (44) |  |
| Treatment |  |  |  | 0.002 |
| • Monotherapy | 41 (45) | 22 (34) | 19 (71) |  |
| • With chemotherapy | 51 (55) | 43 (66) | 8 (29) |  |
| Treatment center |  |  |  | 0.07 |
| • BIDMC | 47 (51) | 29 (45) | 18 (67) |  |
| • VMC | 45 (49) | 36 (55) | 9 (33) |  |
| Median no. of treatment cycles, range | 8 (4-41) | 6 (4-20) | 14 (6-41) | <0.0001 |
| Best response |  |  |  | 0.67 |
| • Progression | 7 (8) | 6 (9) | 1 (4) |  |
| • Clinical benefit | 83 (90) | 57 (88) | 26 (96) |  |
| ○ CR | 15 (16) | 12 (19) | 3 (11) |  |
| ○ PR | 40 (44) | 25 (38) | 15 (56) |  |
| ○ SD | 28 (30) | 20 (31) | 8 (30) |  |
| • Not available | 2 (2) | 2 (3) | - |  |
| Any grade irAE, Yes | 54 (59) | 35 (54) | 19 (70) | 0.17 |
| ≥ Grade 3 irAE, Yes | 28 (30) | 21 (32) | 7 (26) | 0.63 |
| Systemic immunosuppression, Yes | 41 (45) | 29 (45) | 12 (44) | 1.0 |

Abbreviations: ECOG, Eastern Cooperative Oncology Group; PS, performance status; TPS, tumor proportion score; TMB, tumor mutational burden; CR, complete response; PR, partial response; SD, stable disease; irAE, immune-related adverse events.

Median OS was not reached (N.R.) in the Non-Std group and was significantly longer compared to the Std group by univariate analysis [Std: 15.4 (95% C.I.: 9.0 – N.R.) vs. Non-Std: N.R. (95% C.I.: N.R.) months] (**Figure 2A**, **Supplementary Table S3**). Median PFS was also significantly longer in the Non-Std group compared to the Std group by univariate analysis [Std: 7.0 (95% C.I.: 5.1 - 8.8) vs. Non-Std: 23.3 (95% C.I.: 14.6 – N.R.) months] (**Figure 2B**, **Supplementary Table S4**). 6-month landmark analyses continued to show significant differences in both OS [Std: 34.9 (95% C.I.: 15.4 – N.R.) vs. Non-Std: N.R. (95% C.I.: N.R.) months] and PFS [Std: 11.8 (95% C.I.: 8.8 – N.R.) vs. Non-Std: N.R. (95% C.I.: 14.6 - N.R.) months] between the two groups (**Figure 2C-D**). However, after adjustment with multivariable regression (stratified by immune-related adverse events due to its time-variant nature), no significant differences were seen in OS [HR for death, 1.2 (95% C.I.: 0.3 – 4.8)] or PFS [HR for disease progression or death, 2.6 (95% C.I.: 0.7 – 9.6)] between the Non-Std and Std groups (**Table 2 and Table 3**). Swimmer’s plots for patients belonging to the Non-Std group showed that most patients received their first non-standard cycle within 6 months of start of therapy, with most having sustained responses (**Figure 3**). Univariate analyses of OS and PFS by the three predominant indications for non-standard dosing in the Non-Std group compared to the Std group showed statistically significant differences favoring the Non-Std subgroups - except for OS relating to the patient-physician preference (**Supplementary Figure S1**).

**Table 2.** Multivariable adjustment for confounding factors for overall survival by Cox proportional hazards regression model stratified by immune-related adverse events

|  | **HR for death**<br>**(95% C.I.)** | **p** |
| --- | --- | --- |
| Standard vs. Non-standard group | 1.2<br>(0.3 – 4.8) | 0.824 |
| ECOG PS $\geq 2$ vs. 0-1 | 2.4<br>(0.9 – 5.9) | 0.066 |
| Pembrolizumab alone vs. along with chemotherapy | 1.4<br>(0.6 – 3.3) | 0.446 |
| <50% vs. $\geq 50\%$ PD-L1 TPS | 0.8<br>(0.3 – 1.9) | 0.591 |
| No. of treatment cycles | 0.8<br>(0.6 – 0.9) | **0.001** |

Abbreviations: HR, hazard ratio; C.I., confidence intervals; ECOG, Eastern Cooperative Oncology Group; PS, performance status; TPS, tumor proportion score.

**Table 3.** Multivariable adjustment for confounding factors for progression-free survival by Cox proportional hazards regression model stratified by immune-related adverse events

|  | **HR for disease progression or death**<br>**(95% C.I.)** | **p** |
| --- | --- | --- |
| Standard vs. Non-standard group | 2.6<br>(0.7– 9.6) | 0.157 |
| Never vs. current/former smoker | 4.2<br>(1.6 – 11.3) | **0.004** |
| Pembrolizumab alone vs. along with chemotherapy | 2.7<br>(1.2 – 6.2) | **0.016** |
| <50% vs. ≥ 50% PD-L1 TPS | 0.9<br>(0.4 – 2.1) | 0.873 |
| ECOG PS ≥ 2 vs. 0-1 | 0.8<br>(0.4 – 1.9) | 0.700 |
| No. of treatment cycles | 0.7<br>(0.6 – 0.8) | **<0.001** |

Abbreviations: HR, hazard ratio; C.I., confidence intervals; TPS, tumor proportion score; ECOG, Eastern Cooperative Oncology Group; PS, performance status.

**Figure 2.**
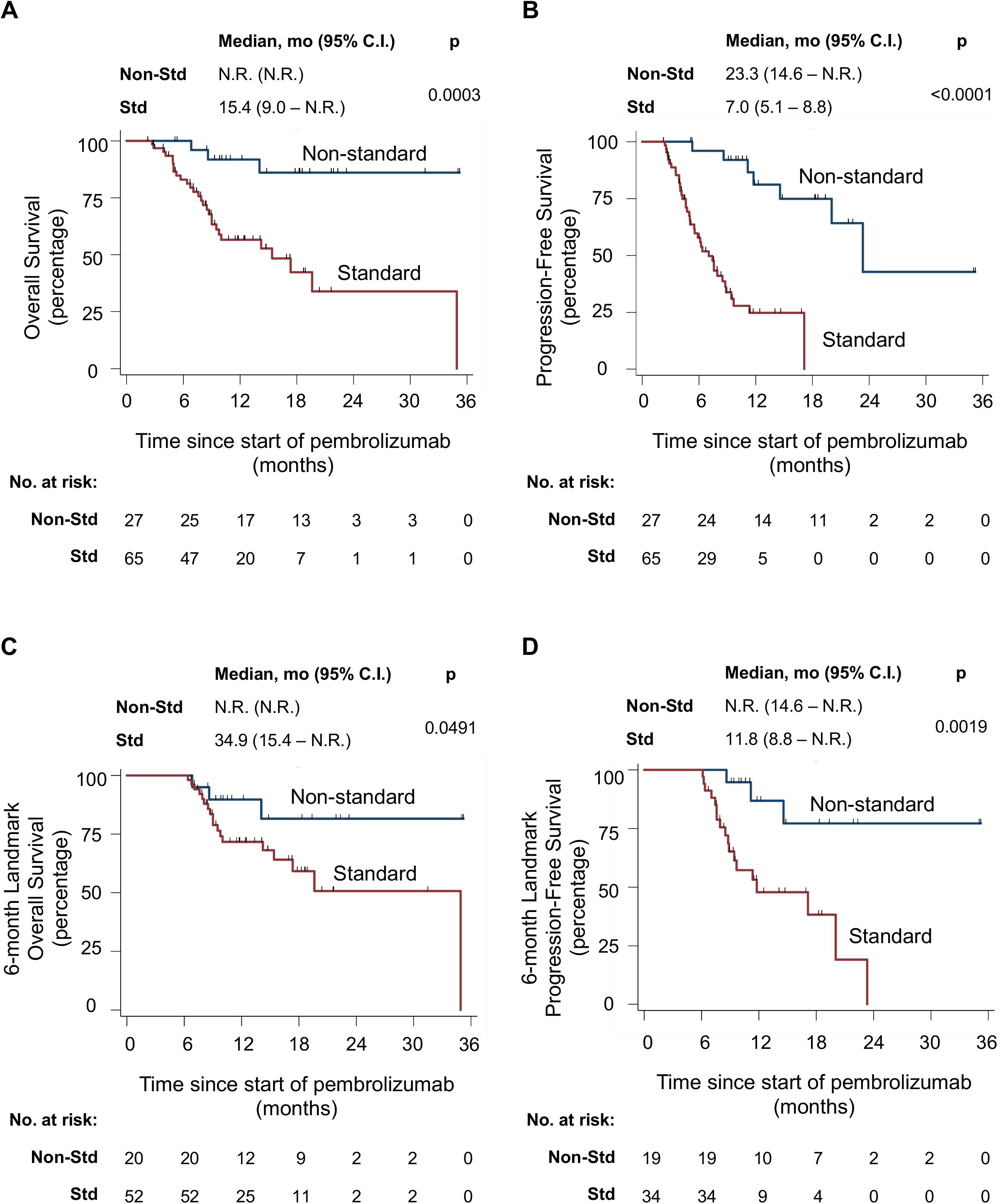
Univariate Kaplan-Meir survival curves in patients with advanced NSCLC belonging to non-standard vs. standard groups for (A) overall survival, (B) progression-free survival, (C) 6-month landmark overall survival, and (D) 6-month landmark progression-free survival. Non-Std, non-standard; Std, Standard; mo, months; C.I., confidence intervals; N.R., not reached.

**Figure 3.**
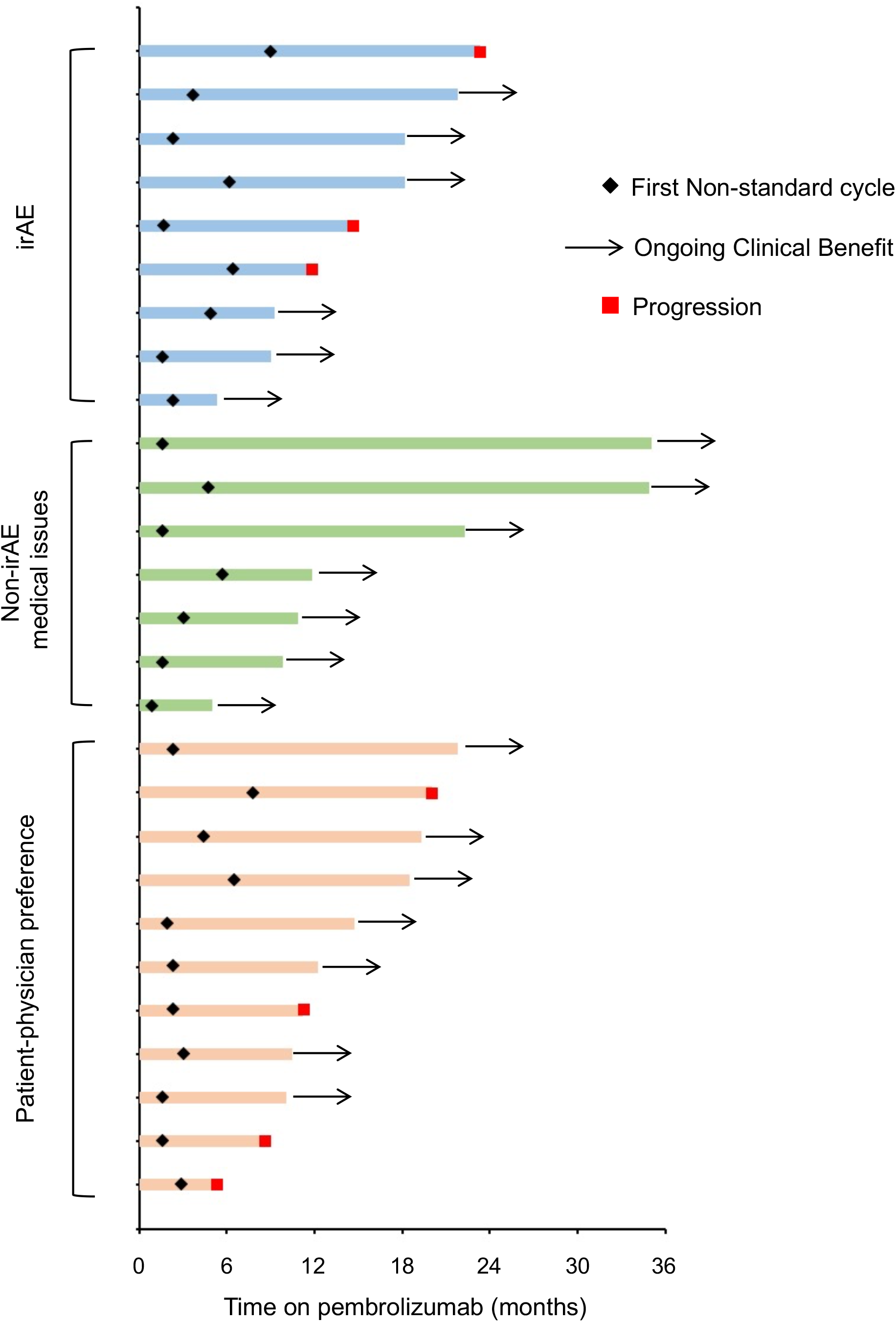
Swimmer’s plot showing time on pembrolizumab treatment after first non-standard (extended or delayed) pembrolizumab cycle in the non-standard group with patients distributed by the indication subgroups. irAE, immune-related adverse events.

## DISCUSSION

We report here the real-world outcomes of patients with advanced NSCLC receiving pembrolizumab-based regimens with extended intervals or treatment delays due to indications commonly encountered in routine clinical practice: irAEs, treatment-unrelated medical issues, and/or individual care preferences. Within the limitations discussed below, these patients had comparable outcomes with those who either received all (or up to 1 delayed cycle of) pembrolizumab at the FDA-approved label of 200 mg every 3 weeks. We acknowledge that our results are hypothesis-generating only, but relevant in an arena where no other well-vetted data exists.

Most early pharmacokinetic and pharmacodynamic studies from phase I clinical trials of pembrolizumab evaluated doses between 2-10 mg/kg every 2-3 weeks.^2,15-17^ These were the basis of a modeling/simulation study which evaluated the exposure-response relationship with extended pembrolizumab dosing interval of 6 weeks, albeit with a higher dose of 400 mg;^13^ this dosing schema was approved by the European Commission and recently by the FDA.^18,19^ Whether extending pembrolizumab dosing intervals while keeping the dose at 200 mg will lead to the same predicted efficacy and safety has not been studied yet. Our data provides rationale for further evaluation of extended dosing intervals of pembrolizumab, particularly in patients with disease response or stabilization after the first four treatment cycles. This may be a more fiscally and logistically viable model, while improving flexibility and patient experience.

Recent pharmacoeconomic analyses comparing alternative dosing strategies of pembrolizumab (including weight-based dosing) to FDA-approved labels have estimated major cost savings for the health system with a personalized approach. ^20,21^ Randomized non-inferiority clinical trials designed with Bayesian methods would be the gold-standard for evaluating these extended dosing regimens in an effective and cost-efficient manner. ^9,22-24^ Alternatively, therapeutic drug monitoring for personalized dosing - as commonly used for antibiotics and immunosuppressive agents - to achieve plasma or serum drug concentrations within a known therapeutic range is another potential strategy that can be employed in prospective studies to minimize financial toxicity from drug and pharmacy costs in this growing population.^9,25^ It would also be prudent to take into account the time-dependent reduction in clearance of immune checkpoint inhibitors in these studies.^26,27^

Limitations of this study include retrospective analysis, small sample size, confounding by indication, exclusion of patients who did not receive at least 4 pembrolizumab-based treatment cycles, and inclusion of patients treated only at tertiary academic cancer centers. These results are not applicable to patients whose disease progresses earlier in the treatment course and those being treated in other practice settings. Even though we employed 6-month landmark survival analysis and multivariable regression to account for the guaranteed time bias and confounding variables, respectively, these biases persist. These findings require vetting in a large prospective manner. Moreover, it is not possible to draw any definitive conclusions when comparing the three predominant subgroups of the Non-Std group to the Std group due to the small sample sizes. Tumor mutation burden was not included in the final adjusted model, as it was available for only approximately 50% of the patients and was not measured with a uniform assay.

## CONCLUSIONS

To the best of our knowledge, this is the first study to describe outcomes of patients with advanced NSCLC receiving pembrolizumab-based regimens at extended intervals due to real-world situations commonly faced in routine clinical practice and unprecedented circumstances such as COVID-19 pandemic. Within the limitations described above, our study provides rationale for prospectively evaluating the administration of the lowest and least frequent efficacious dose of pembrolizumab, particularly for patients with demonstrated disease stability or response for the first 3-6 months.

Clinical Practice Points:

- The most cost-effective administration frequency of pembrolizumab in advanced non-small cell lung cancer (NSCLC) has not been evaluated in clinical trials. Based on a modeling/simulation study, the dosing schedule of pembrolizumab at 400 mg every 6 weeks has been approved by the European Commission and U.S. FDA.
- In this multicenter retrospective cohort study, we found that a significant proportion of patients with advanced NSCLC receive pembrolizumab-based regimens with extended intervals or delays in routine clinical practice due to immune-related adverse events, medical issues and patient-physician preferences.
- We found that these treatment delays or extended dosing intervals were not associated with worse outcomes after multivariable adjustment for confounding factors in the patients with advanced NSCLC who had received at least 4 cycles of pembrolizumab-based regimens.
- To the best of our knowledge, this is the first study to describe outcomes of patients with advanced NSCLC receiving pembrolizumab-based regimens at extended intervals due to real-world situations commonly faced in routine clinical practice.
- Prospective evaluation of alternative dosing strategies in randomized non-inferiority clinical trials, with attention to time-dependent reduction in clearance of pembrolizumab and potential incorporation of personalized dosing with therapeutic drug monitoring is warranted.
- Alternative dosing strategies may provide a more fiscally and logistically viable model, while improving flexibility and patient experience.

## Data Availability

Deidentified data will be available upon reasonable request to the corresponding authors.

## Acknowledgements

Preliminary findings of this work have been reported at the 2019 ASCO annual meeting, Chicago, USA and the 2019 IASLC World Conference on Lung Cancer, Barcelona, Spain.

## Author Contributions

**Kartik Sehgal:** Conceptualization, Data curation, Formal analysis, Methodology, Software, Visualization, Roles/Writing - original draft. **Anushi Bulumulle:** Investigation, Writing - review & editing. **Heather Brody:** Investigation, Writing - review & editing. **Ritu R. Gill:** Investigation, Methodology, Writing - review & editing. **Shravanti Macherla:** Investigation, Writing - review & editing. **Aleksandra Qilleria:** Investigation, Writing - review & editing. **Danielle C. McDonald:** Investigation, Writing - review & editing. **Cynthia R. Cherry:** Investigation, Writing - review & editing. **Meghan Shea:** Investigation, Writing - review & editing. **Mark S. Huberman:** Investigation, Writing - review & editing. **Paul A. VanderLaan:** Investigation, Writing - review & editing. **Glen J. Weiss:** Investigation, Methodology, Writing - review & editing. **Paul R. Walker:** Project administration, Project administration, Resources, Supervision, Writing - review & editing. **Daniel B. Costa:** Conceptualization, Funding acquisition, Project administration, Resources, Supervision, Roles/Writing - original draft. **Deepa Rangachari:** Conceptualization, Methodology, Project administration, Resources, Supervision, Roles/Writing - original draft.

## Footnotes

1 Present address: UNUM Therapeutics Inc., 200 Cambridge Park Drive, Suite 3100, Cambridge, MA 02140, USA

## 2 Abbreviations

BIDMC: Beth Israel Deaconess Medical Center, Non-Std: Non-standard, N.R.: Not reached, Std: Standard, VMC: Vidant Medical Center

## MicroAbstract

The most cost-effective administration frequency of pembrolizumab in advanced non-small cell lung cancer (NSCLC) is unknown. We found that a significant proportion of these patients receive pembrolizumab-based regimens with extended intervals or delays in routine practice, with similar outcomes to those on label-specified 3-week interval treatments. Prospective evaluation of alternative dosing strategies is warranted to develop a more fiscally viable and patient-centered model.

---

**Supplementary Figure S1**. Univariate survival curves in patients with advanced NSCLC belonging to the standard group versus subgroups of the non-standard group for (A) overall survival and (B) progression-free survival.

**Supplementary Table S1**. Patient characteristics of the screened population.

**Supplementary Table S2**. Reasons for delays or extensions of pembrolizumab-based cycles in the non-standard group.

**Supplementary Table S3**. Univariate analysis of overall survival by Cox proportional hazards regression model.

**Supplementary Table S4**. Univariate analysis of progression-free survival by Cox proportional hazards regression model.

